# Preschool Teacher Education and Child-Teacher ratio in Relation to Children’s Health: A Large-Scale Study of Preschoolers in Stockholm Sweden

**DOI:** 10.1101/2025.11.20.25340638

**Authors:** Charlotte Wilén, Pablo Campos Garzón, Jairo Hidalgo Migueles, Pontus Henriksson, Chu Chen, Per Tynelius, Daniel Berglind, Viktor H. Ahlqvist

**Author notes:** **Correspondence:** Charlotte Wilén. Shared senior author (Daniel Berglind, Viktor H. Ahlqvist).

## Abstract

**Objective:** To examine whether teacher education levels and child–teacher ratios are associated with children’s physical activity, psychosocial functioning, and BMI in a large, population-based sample of preschoolers in Stockholm, Sweden.

**Study design:** In a cross-sectional study of 124 public preschools, we analyzed data from two measurement occasions (2020 and 2021) for 3,302 children with accelerometer data, 2,666 with Strengths and Difficulties Questionnaire (SDQ) scores, and 3,518 with measured BMI. Teacher education and child–teacher ratios were obtained from the Swedish National Agency for Education. Mixed-effects regression models accounted for clustering and adjusted for demographic and neighborhood characteristics.

**Results:** Across preschools, an average of 31.8% of full-time teachers had a teaching degree, and the mean child–teacher ratio was 5.1 (SD 0.7). No strong associations were observed between teacher education levels or child–teacher ratios and children’s moderate-to-vigorous physical activity (MVPA), inactivity, SDQ, or BMI. Compared with the median, attending preschools with teacher education levels above the 75^th^ percentile was associated with −0.22 minutes (95% CI: −0.84 to 0.40) of daily MVPA, and the 25^th^ percentile with −0.04 minutes (95% CI: −0.77 to 0.69). Similarly, increasing the child–teacher ratio from 5.0 to 5.5 children per teacher corresponded to −0.45 minutes (95% CI: −1.17 to 0.28) of daily MVPA, with negligible differences in SDQ and BMI z-scores.

**Conclusions:** In this large, representative sample of Stockholm preschoolers, teacher education levels and child–teacher ratios were not associated with children’s physical activity, psychosocial functioning, or BMI, suggesting that factors beyond staffing characteristics may play a more central role in shaping children’s health in preschool settings.

## Introduction

Preschools provide a natural and strategically important setting for promoting children’s health and development. In many countries, including Sweden, a large proportion of children attend preschool (1, 2), and research has examined factors shaping children’s behavior and long-term health outcomes in these settings (3, 4). Much of this work has focused on modifying the physical environment, such as upgrading play areas, or implementing policy-driven interventions (5-8), with mixed results and often inconclusive findings (9-11).

One factor that has received less empirical attention, despite emphasis by teacher unions and regulatory agencies, is the role of staff qualifications and child–teacher ratios. Higher ratios of teachers to children and a greater proportion of formally trained staff may support child development by enabling more individualized attention, closer supervision, and expanded opportunities for structured physical and social engagement. Conversely, excessive adult oversight or rigidly applied pedagogical frameworks could suppress spontaneous activity or disrupt unstructured peer interactions (12-14). Notably, preschool staff are often conceptualized as passive facilitators of externally led interventions rather than as active determinants of the preschool environment (15).

Despite their potential influence, few studies have directly examined how teacher staffing and qualifications affect children’s health (12-14). Existing research has yielded mixed findings, with generally modest associations, and has often been limited by small sample sizes, lack of standardized measures, and a focus on social or developmental outcomes rather than objective health indicators (12-14). Thus, the relationship between staffing policies and children’s health remains uncertain.

In this study, we leverage comprehensive, population-based data from preschools in Stockholm, Sweden, to examine how staff qualifications and child-teacher ratios are associated with children’s physical activity, psychosocial functioning, and body mass index (BMI). By using objective and standardized measures across a large and socioeconomically diverse population, this work aims to inform early childhood policy at a time when promoting health within educational settings is an increasingly urgent priority.

## Methods

Ethical approval was obtained from the Swedish Ethical Review Authority (DNR 2020-03002), and written informed consent was obtained from all participants caregivers. The study was conducted in accordance with the Strengthening the Reporting of Observational Studies in Epidemiology (STROBE) guidelines (16).

### Derivation of study population

This study utilizes data from the Increasing Children’s Physical Activity by Policy (CAP) study, which has been described in detail elsewhere (17). In short, CAP is an RCT assessing an intervention to promote physical activity, comprising an intervention group and a control group. Here, we use the baseline and follow-up as a cross-sectional analytical sample, and adjust for the period of measure. To ensure a representative sample reflecting the socioeconomic diversity of Stockholm, all public preschools across Region Stockholm’s 13 districts were randomly assigned to an invitation sequence clustered by district. Preschools were then invited in this order until at least 30% of all public preschools in each district agreed to take part in the study. All children attending the participating preschools were invited to participate in the study, with parental informed consent required for inclusion. A total of 124 preschools and 3,751 children participated in the CAP study, with baseline data collected during spring 2020 and follow-up during autumn 2021.

### Teacher education and child-teacher ratio

Data on teacher education and child-teacher ratio were obtained from publicly available records kept by the Swedish National Agency for Education, the government authority responsible for official statistics on the school and childcare systems. Reporting to the agency is mandatory for institutions conducting educational activities. The data were extracted from October 2020 and October 2021, to best correspond with the time at which the children’s parameters were collected. One school could not be located in the 2021 records; therefore, data from 2020 were used for both measurement points for that school (excluding or including this school did not materially affect any results).

Teacher education was defined as the proportion (%) of full-time positions with a preschool teaching degree, rounded to the closest integer. Child-teacher ratio was defined as the number of enrolled children per full-time position, regardless of the educational background or certification of the staff member. A full-time position was defined as preschool staff, regardless of educational background, who were employed for a minimum of one month and worked directly with the children (i.e., not purely administrative). Percentages of part-time employment were summed up, meaning that two staff members working half-time count as one full-time position. On-call and hourly-paid staff were not included.

### Participants and Inclusion Criteria

To maximize the number of observations in the study, we applied general inclusion criteria and additional outcome-specific criteria. General inclusion criteria required participants to have valid data on age, sex and neighborhood socioeconomic index. As for the outcome-specific criteria, three separate analytical samples were defined: 1) participants with valid accelerometry data (10% of the total children from CAP sample excluded); 2) participants with valid data on psychosocial functioning (27% of the total children from CAP sample excluded) and 3) participants with valid BMI data (5% of the total children from CAP sample excluded) (Fig. 1). All three outcomes were measured at the data collection occasions both in 2020 and 2021.

**Fig. 1.**
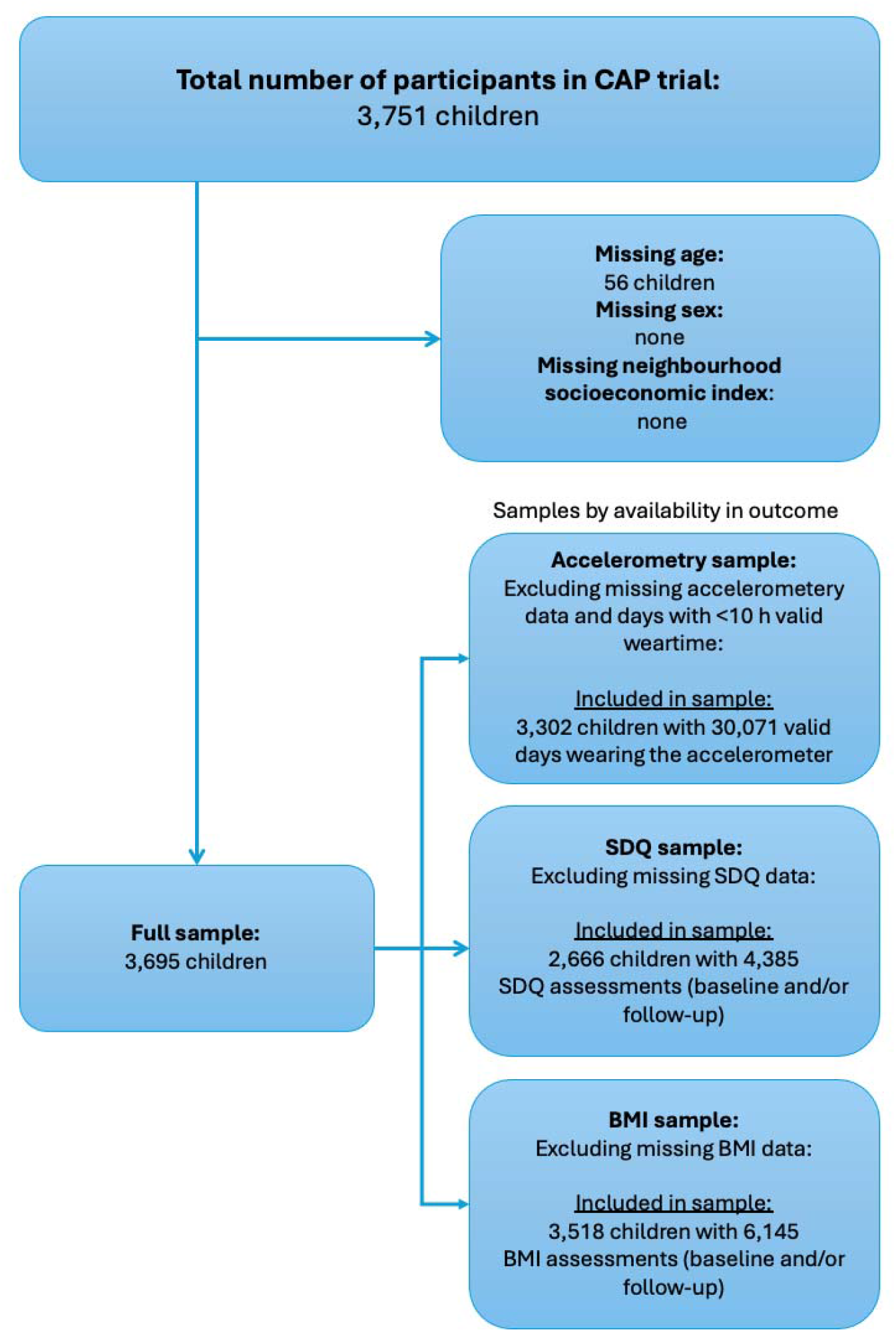
Derivation of study samples. A day or assessment could come from either autumn 2020, spring 2021, or both. Thus, since children were instructed to wear the accelerometer for one week per occasion, a child could contribute multiple measurements per occasion for physical activity, but only one measurement per occasion for SDQ and BMI.

### Physical activity

Physical activity was measured using GT3X+ accelerometers, which the children were instructed to wear on their non-dominant wrist continuously, day and night, for one week at each measurement point (one week in spring and one week in autumn), except during water-based activities. The accelerometers were set to a sampling frequency of 30 Hz, and data were analyzed in 5-second epochs, which is appropriate for capturing the sporadic movement patterns typical of preschool-aged children (18). Accelerometer data were considered valid if the child wore the device for more than 10 hours per day on any measurement point (baseline in autumn 2020 or follow-up in spring 2021). Details on accelerometer data processing have been published elsewhere.

This study examined full-day and -week moderate-to-vigorous physical activity (MVPA) and time being inactive, classified according to the cut-off values established by Hildebrand et al. (19, 20). Further, meeting World Health Organization’s (WHO) recommendations of at least 60 minutes of MVPA each day (21) was examined as an outcome.

### Psychosocial functioning

Children’s psychosocial functioning was assessed using the validated Strengths and Difficulties Questionnaire (SDQ), completed by parents (22). The questionnaire includes five items across four scales: emotional symptoms, conduct problems, hyperactivity/inattention, and peer relationship problems. Scores of each scale were calculated by summing the responses within each scale, with higher scores indicating greater psychosocial difficulties. A total difficulties score was then derived by summing the four scale’s scores. SDQ z-scores were calculated by standardizing each observation’s total difficulties score using the mean and standard deviation of the total scores across all observations.

### Body Mass Index

Children’s height and weight were measured using stadiometers and validated scales. Each measurement was recorded to the nearest 0.1 unit and taken twice, to obtain an average. BMI was then calculated as weight in kilograms divided by height in meters squared (kg/m^2^). BMI z-scores were calculated by internally standardizing BMI within each sex and age groups (per 0.5 year). For the descriptive characteristics, weight status was categorized as having normal weight, overweight, or obesity according to the criteria developed by Cole et al (23).

### Covariates

Age and sex were derived from the child’s social security number reported by the parents upon giving informed consent. Age was rounded to the closest half-year. As season (winter or summer) may influence the outcomes, especially physical activity (24), the period in which the measurement was taken (autumn 2020 or spring 2021) was included as a covariate in all analyses. The neighborhood socioeconomic index, developed by Region Stockholm using 2019 data, was used to characterize the socioeconomic context of each school area (25). The index is based on several indicators, including rates of unemployment, receipt of financial assistance, levels of education below the secondary level, prevalence of single-parent households, proportion of foreign-born residents, median income, and average living space. Each child was assigned the socioeconomic index corresponding to the location of their preschool.

### Statistical analysis

We first described the three study samples using appropriate summary statistics, including measures of central tendency and dispersion.

To evaluate associations between teacher education and child-teacher ratio and continuous measures of children’s health, we employed mixed-effects linear and logistic regression models. The unit of analysis was the measurement occasion. Each child contributed one or more observations, depending on the availability of valid data; a child could contribute data from either or both measurement waves in the CAP study, and for physical activity, for repeated days within each measurement wave. To account for within-child and within-school clustering, we specified three-level hierarchical models with random intercepts at the child and school levels, with children nested within schools. To quantify the proportion of the total variability attributable to each level, we estimated the intraclass correlation coefficient (ICC) for each outcome using null models.

To allow for potential non-linear associations, the teacher education and child-teacher ratio were modelled using restricted cubic splines with four knots positioned at the 5^th^, 35^th^, 65^th^ and 95^th^ percentiles, following Harrell’s recommendations (26). Due to limited data beyond the 5^th^ and 95^th^ percentiles (Supplementary material, fig. S1 A & B), the cubic spline figures were restricted to this range to reduce the risk of misleading interpretations.

The results are reported on the absolute scale (marginal means for continuous outcomes or marginal prevalences for binary outcomes) and on the relative scale (mean differences or odds ratios for binary outcomes) with 95% confidence intervals. Relative estimates are expressed as contrasts comparing the 25^th^ and 75^th^ percentiles to the median (50^th^ percentile) of the exposure distribution (Supplementary material, fig. S1 A & B) from the underlying restricted cubic spline model. All exposure values at the different percentiles were estimated based on the full sample (Fig. 1). Percentile positions were consistent across all subsamples (Supplementary material, Table S1). Exact percentile values for all categories are provided in Supplementary material, Table S1.

To assess potential interactions between teacher education and the child–teacher ratio, we examined associations across combinations of high/low teacher education and high/low child– teacher ratios.

All models were adjusted for child age, sex, measurement period (autumn 2020 vs. spring 2021) and neighborhood socioeconomic index. Since we expect some correlation between teacher education and the child–teacher ratio, models including teacher education were adjusted for child–teacher ratio and vice versa. Analyses were conducted using Stata version 19.0.

### Sensitivity analyses

Several sensitivity analyses were conducted: (i) stratification by age (<4 vs. ≥4 years) and sex; (ii) restricting accelerometer data to school hours only, with a school day considered valid if ≥5 hours of data were recorded; (iii) excluding children with fewer than three valid accelerometer days; (iv) stratification by measurement period (autumn 2020 vs. spring 2021); and (v) stratifying by allocation to control or intervention preschool.

## Results

### Population characteristics

The analytic samples included 3,302 children for physical activity, 2,666 for SDQ and 3,518 for BMI (**Table 1**). The corresponding number of measurements was 30,071 days for accelerometry, 4,385 for SDQ responses, and 6,145 for BMI measurements. Across preschools, the mean proportion of full-time teachers with a teaching degree was 31.8%, and the mean child-to-teacher ratio was 5.1 (SD 0.7). These staffing characteristics were similar across the three outcome samples.

**Table 1.** Descriptive characteristics of the children included at each measurement point within the three overlapping CAP samples: accelerometer-measured physical activity, parent-reported Strengths and Difficulties Questionnaire (SDQ), and measured body mass index (BMI).

|  | Sample 1:<br>Accelerometry<br>sample | Sample 2: SDQ<br>sample | Sample 3: BMI<br>sample |
| --- | --- | --- | --- |
| **Participant data** |  |  |  |
| **N number of participants** | N=3,302 | N=2,666 | N=3,518 |
| **Demographic information** |  |  |  |
| Year of measurement |  |  |  |
| Only 2020 | 639 (19.4%) | 595 (22.3%) | 423 (12.0%) |
| Only 2021 | 441 (13.4%) | 352 (13.2%) | 468 (13.3%) |
| Both years | 2,222 (67.3%) | 1,719 (64.5%) | 2,627 (74.7%) |
| Girls, N(%) | 1,604 (48.6%) | 1,295 (48.6%) | 1,702 (48.4%) |
| Age, Mean (SD) | 4.7 (0.9) | 4.7 (0.9) | 4.7 (0.9) |
| Neighborhood socioeconomic index, N(%) |  |  |  |
| Low (Q1) | 1,164 (35.3%) | 868 (32.6%) | 1,257 (35.7%) |
| Medium (Q2) | 1,108 (33.6%) | 914 (34.3%) | 1,159 (32.9%) |
| High (Q3) | 1,030 (31.2%) | 884 (33.2%) | 1,102 (31.3%) |
| **Exposure and outcome data** |  |  |  |
| **K number of measurement points** | K=30,071 | K=4,385 | K=6,145 |
| **Teacher Education and Child-Teacher ratio** |  |  |  |
| Proportion (%) of full-time positions with a preschool teaching degree, Mean (SD) | 31.8 (7.6) | 31.7 (7.5) | 31.8 (7.7) |
| Number of enrolled children per full-time position, Mean (SD) | 5.1 (0.7) | 5.1 (0.7) | 5.1 (0.7) |
| **Physical activity/day (accelerometer)** |  |  |  |
| Steps (count), Mean (SD) | 13176.1 (4269.1) | 13191.2 (4265.6) | 13284.2 (4333.3) |
| Sedentary time (min), Mean (SD) | 432.3 (112.8) | 421.6 (117.1) | 417.8 (120.7) |
| Light physical activity (min), Mean (SD) | 309.2 (66.9) | 303.2 (69.3) | 299.0 (72.4) |
| Moderate to vigorous physical activity (min), Mean (SD) | 61.2 (29.2) | 60.2 (28.9) | 59.6 (28.9) |
| Meets MVPA recommendations, $\geq 60$ min/day, N (%) | 45.7% | 43.7% | 42.7% |
| **SDQ and BMI measures** |  |  |  |
| Parent-reported SDQ-scores, Mean (SD) | 6.2 (4.2) | 6.3 (4.3) | 6.2 (4.2) |
| Parent-reported SDQ z-scores, Mean (SD) | -0.0 (1.0) | 0.0 (1.0) | 0.0 (1.0) |
| BMI, % |  |  |  |
| Normal | 78.9% | 76.6% | 80.7% |
| Overweight or obese | 18.1% | 16.9% | 19.3% |
| Missing | 3.1% | 6.4% | . |
| BMI z-scores, Mean (SD) | -0.0 (1.0) | -0.0 (1.0) | 0.0 (1.0) |

Intraclass correlation coefficients showed that only a small proportion of variance was explained by differences between preschools (MVPA 2.4%, inactivity 2.5%, SDQ 1.8%, BMI 3.3%), whereas most variability reflected differences within children, within preschools (MVPA 39.0%, inactivity 23.0%, SDQ 73.0%, BMI 85.2%).

### Preschool teacher education and child-teacher ratio and children’s health

The proportion of educated teachers did not have a strong association with physical activity **(Figure 2 & Supplementary material, Table S2)**; compared to the median, the 75^th^ percentile of and 25^th^ percentile were associated with -0.22 (95% CI -0.84 to 0.40) and -0.04 (95% CI -0.77 to 0.69) minutes MVPA per day on average (**Figure 3 & Supplementary material, Table S2**). The same small magnitude was observed for inactivity, albeit the association was statistically significant, with those with a higher proportion of teacher education having slightly more inactivity (3.19 minutes more than the median; 95% CI: 0.67 to 5.70), and those with less educated teacher having less inactivity (4.04 minutes less than the median; 95% CI: -7.05 to -1.02). The same was true for SDQ and BMI – there were only differences at the second decimal of the Z-scores of these across the distribution of teacher education levels.

**Fig. 2.**
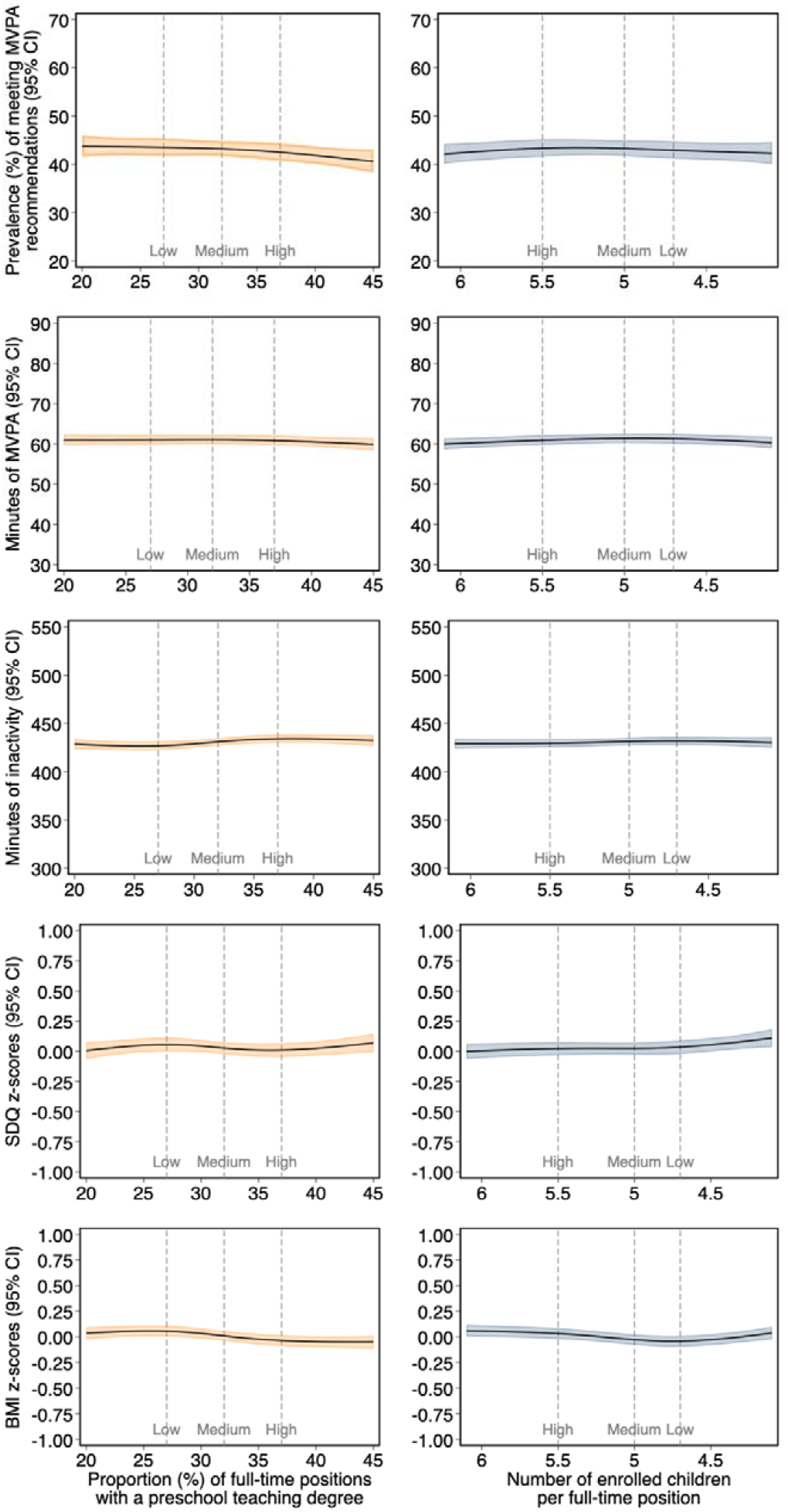
Restricted cubic spline models displaying the associations between both exposures and all outcomes. Knots were placed at the 5^th^, 35^th^, 65^th^ and 95^th^ percentiles, and the splines were restricted to the range between the 5^th^ and 95^th^ percentiles (Supplementary material, fig. S1 A & B). Adjustments were made for child age, sex, measurement point (autumn 2020 spring 2021) and neighborhood socioeconomic index. Models including teacher education were adjusted for child–teacher ratio and vice versa.

**Fig. 3.**
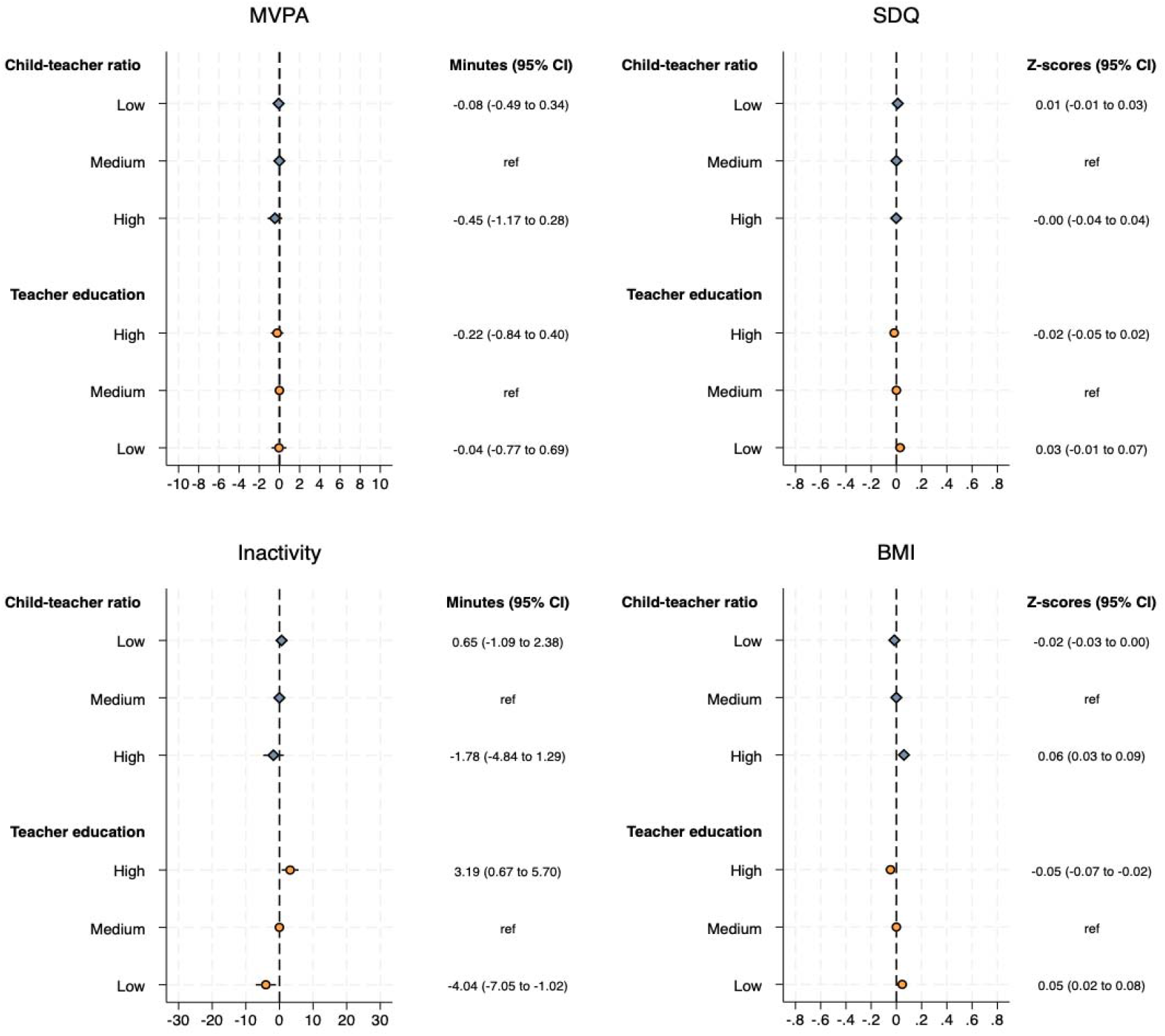
Forest plots showing associations between exposures and outcomes at the 25^th^, 50^th^, and 75^th^ exposure percentiles from the spline models. Results are presented on a relative scale, using the 50^th^ percentile as the reference category. Adjustments were made for child age, sex, measurement point (autumn 2020 spring 2021) and neighborhood socioeconomic index. Models including teacher education were adjusted for child–teacher ratio and vice versa.

Similarly, the child-teacher ratio demonstrated no strong association with physical activity, SDQ, or BMI (**Figure 2 & Supplementary material, Table S2**). For example, having 5.5 as opposed to 5.0 children per teacher was associated with a difference of -0.45 minutes (95% CI: -1.17 to 0.28) MVPA per day, and a difference of 0.00 (95% CI: -0.04 to 0.04) units in SDQ Z-score, and a difference of 0.06 (95% CI: 0.03 to 0.09) units BMI z-score (**Figure 3 & Supplementary material, Table S2**).

### The interaction of different combinations of staffing profiles

Considering teacher education levels and child–teacher ratios jointly led to similar conclusions, with no major differences observed. For example, the hypothesized “worst-case” scenario, low proportion of educated teachers and high child–teacher ratio, was associated with a small decrease in daily MVPA (-1.48 minutes; 95% CI, -2.47 to -0.48) compared with a reference group with a mid-level ratio (**Figure 4**). For inactivity, the combination of the highest proportion of educated teachers and the lowest child–teacher ratio corresponded to a slight increase in daily inactivity (3.60 minutes; 95% CI, 1.12 to 6.08), whereas high teacher education and high child–teacher ratio was associated with a modest reduction in inactivity (-5.88 minutes; 95% CI, -9.76 to -2.01). No evidence of associations was observed for any combinations of exposures with children’s SDQ or BMI z-scores.

**Fig. 4.**
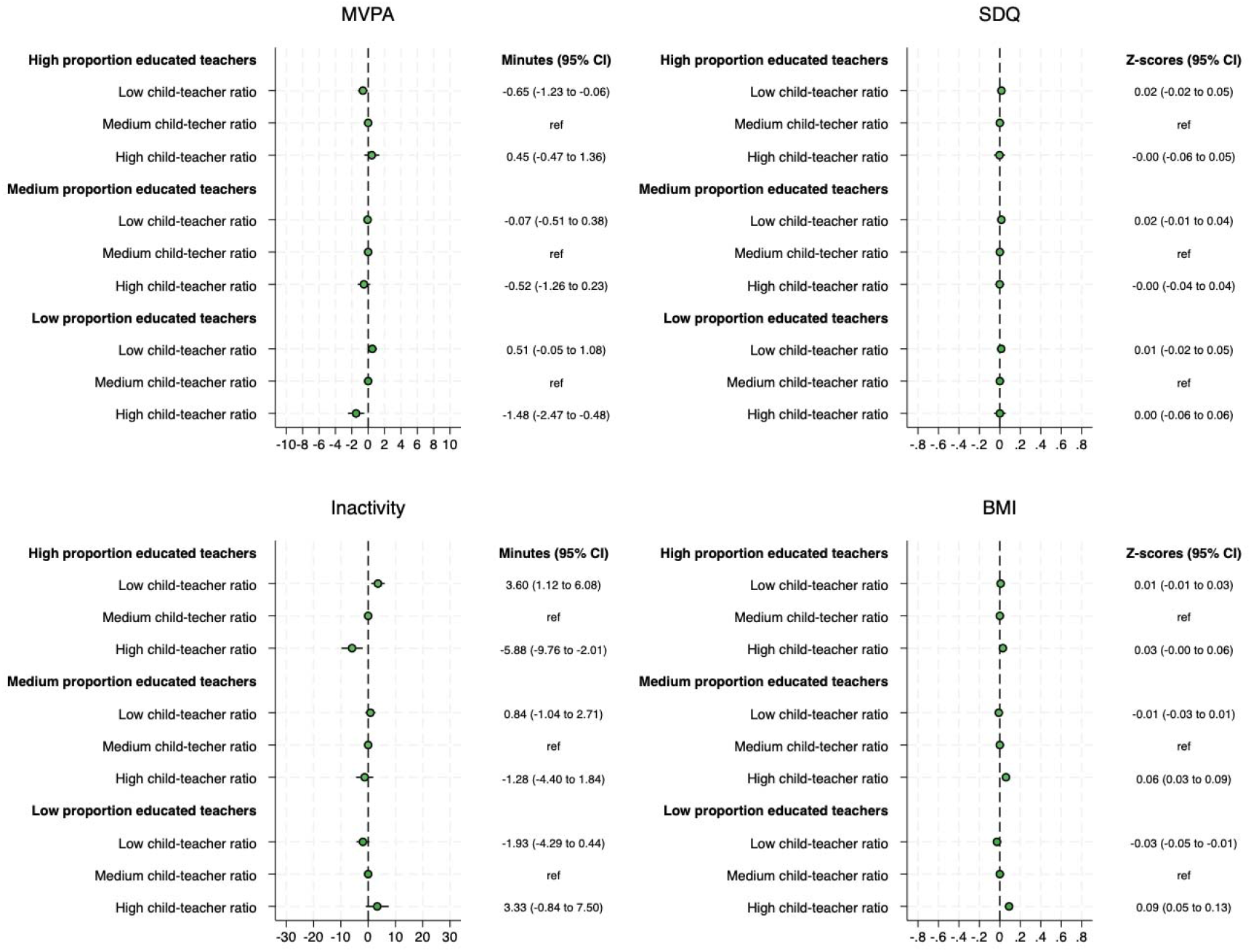
Forest plots showing associations between outcomes and all combinations of exposures at the 25^th^, 50^th^, and 75^th^ percentiles from the spline models. Results are presented on a relative scale, using the 50^th^ percentile as the reference category. Adjustments were made for child age, sex, measurement point (autumn 2020 spring 2021) and neighborhood socioeconomic index.

### Sensitivity analyses

Sensitivity analyses were largely consistent with the main findings. No differences were observed in physical activity when restricted to school hours (**Supplementary material**, Figure S2), by sex or age (**Supplementary material**, Figure S3), or after excluding children with fewer than three valid school days (**Supplementary material**, Figure S5). The only exception was the measurement period (autumn 2020 vs. spring 2021), where at the extreme ends of the distribution of teacher education, MVPA appeared higher during the spring of 2021 (**Supplementary material**, Figure S4). Specifically, during spring 2021, children in preschools with 35% educated teachers averaged 64.3 minutes per day in MVPA (95% CI, 61.3–67.3), whereas those at 45% educated teachers averaged 76.3 minutes (95% CI, 71.1–81.5).

## Discussion

In this large-scale study of a representative sample of preschool children in Stockholm, we found no meaningful association between teacher education levels or child-to-teacher ratios and children’s physical activity, psychosocial functioning, or BMI. Moreover, even when considering these factors jointly, potentially reflecting an ideal scenario with both highly educated teachers and low child-to-teacher ratios, children’s health outcomes were not measurably better. Sensitivity analysis revealed a difference in MVPA between different proportions of educated teachers. However, this pattern was limited to the extreme top of the distribution of proportion of educated teachers (i.e., it was not a linear trend across the entire distribution), and was only observed in autumn.

### Comparison with previous studies

Previous systematic reviews and meta-analyses of the relatively small body of research on child-to-teacher ratios have primarily examined social behaviors, language development, and patterns of interaction between children and adults (12-14). Although we were not able to assess these outcomes, we found no indication that differences in child-teacher ratios translated into measurable differences in children’s health across preschools. Even for the outcomes previously studied (12, 13), reported associations have generally been modest, and our findings are broadly consistent with this lack of association. Interestingly, we could not see that the findings were materially different for physical activity even if only focusing on the within-school hours window, where one perhaps would expect any results to be most evident.

By contrast, a systematic review of teacher education and the quality of early childhood learning environments (14) reported a positive association between higher levels of teacher education and classroom quality. While we found no evidence that improvements in classroom quality, as indicated by teacher education and child–teacher ratio, translate into differences in physical activity, psychosocial functioning, or BMI, it is notable that we observed no strong associations whatsoever. One might have expected that higher levels of teacher qualifications would be reflected in at least some of the health outcomes examined in this study, but instead, we found no evidence of any meaningful association.

### Implications of findings

Although our findings do not support an association between teacher education levels or child-teacher ratios and children’s physical activity, psychosocial functioning, or BMI, several considerations may help interpret these null results. First, preschool teacher education in Sweden includes little formal training in physical activity promotion, which may help explain the absence of observed effects on activity levels. Incorporating a stronger focus on physical activity into teacher training could therefore represent an opportunity to support healthier movement patterns among children. Second, staff characteristics such as educational background or child–staff ratios may simply play a smaller role than often assumed. Opportunities for active play may depend more on the presence of peers or on broader structural and policy factors within preschools than on staffing profiles per se. Nevertheless, taken together, the weak associations observed suggest that policymakers might achieve greater impact by prioritizing environmental or programmatic interventions rather than focusing solely on staffing policies.

### Strengths and limitations

A key strength of this study is the large, population-based sample that includes children from all socioeconomic strata in the Stockholm area. Physical activity was assessed using objective measurements, which are considered superior to self-reported data. In addition, information on teacher qualifications and staffing was obtained from standardized external records, which may provide a more reliable measure than parental or teacher reports.

Several limitations should be acknowledged. First, because the study population was drawn from Stockholm, the findings may not be generalizable to less urban or rural settings. Second, although teacher data were obtained from the Swedish National Agency for Education, data that are reviewed and used for policy decisions, we were unable to validate these measures through direct observations at the preschool level. We also lacked information on hourly paid or on-call staff, who are not included in these estimates but may vary across preschools. Third, as with all observational studies, confounding cannot be ruled out, although it seems unlikely to explain the null findings. Finally, we were unable to account for curricular differences between preschools, such as variation in outdoor time or the extent of structured physical activity. Despite these limitations, the study provides robust population-level evidence on the lack of association between preschool staffing profiles and children’s health outcomes.

### Conclusions

In this large, population-based study of preschool children in Stockholm, we found no evidence that teacher qualification levels or child–teacher ratios were associated with differences in children’s physical activity, psychosocial functioning, or BMI. Even under favorable staffing profiles, no consistent benefits were observed. The limited associations underscores that these structural factors may play only a minor role in shaping health behaviors in early childhood, compared with other environmental or policy-related influences. Future research should examine the interplay between preschool environments, teacher practices, and broader policy frameworks, and assess whether targeted training of educators in physical activity promotion could yield more consistent effects.

## Data Availability

The data underlying this article cannot be shared publicly due to Swedish legislation as well as the privacy of individuals that participated in the study.

## Abbreviations

BMI: Body Mass Index
CAP: Children’s Physical Activity by Policy
MVPA: Moderate-to-Vigorous Physical Activity
SDQ: Strengths and Difficulties Questionnaire
WHO: World Health Organization

## Declarations

### Competing interests

The authors declare no competing interests.

### Funding

Open access funding provided by Karolinska Institutet. This work was supported by the Health and Medical Care Administration (Hälsooch Sjukvårdsförvaltningen, Region Stockholm) and by grants to the senior author/PI DB from: (i) the Swedish Cancer Society (Cancerfonden) and (ii) Swedish Research Council for Sport Science (Centrum för Idrottsforskning). The funders had no role in study design, data collection, analysis or interpretation of data, writing the manuscript or decision to publish.

### Ethics approval and consent to participate

Ethical approval was obtained by the Swedish Ethical Review Authority (DNR: 2020-03002). Informed consent was obtained from the caregivers to all participating children. Data were pseudo anonymized for all analyses and were accessible only by authorized personnel.

### Author’s contributions

CW: data analysis and writing, VHA: data analysis, data interpretation, writing, supervision PT: data analysis, data interpretation, editing, PH/CC/JHM/PCG: data interpretation, editing, DB: study design (PI), data interpretation, editing, supervision.

## Acknowledgements

The authors would like to acknowledge all the participating children, parents and preschool staff.

## Notes

### Competing Interest Statement

The authors have declared no competing interest.

### Clinical Trial

ClinicalTrials.gov (NCT04569578).

### Author Declarations

Ethical approval was obtained by the Swedish Ethical Review Authority (DNR: 2020-03002).

