## Supplementary materials for "Preschool Teacher Education and Child-Teacher ratio in Relation to Children’s Health: A Large-Scale Study of Preschoolers in Stockholm Sweden"

### Supplementary material

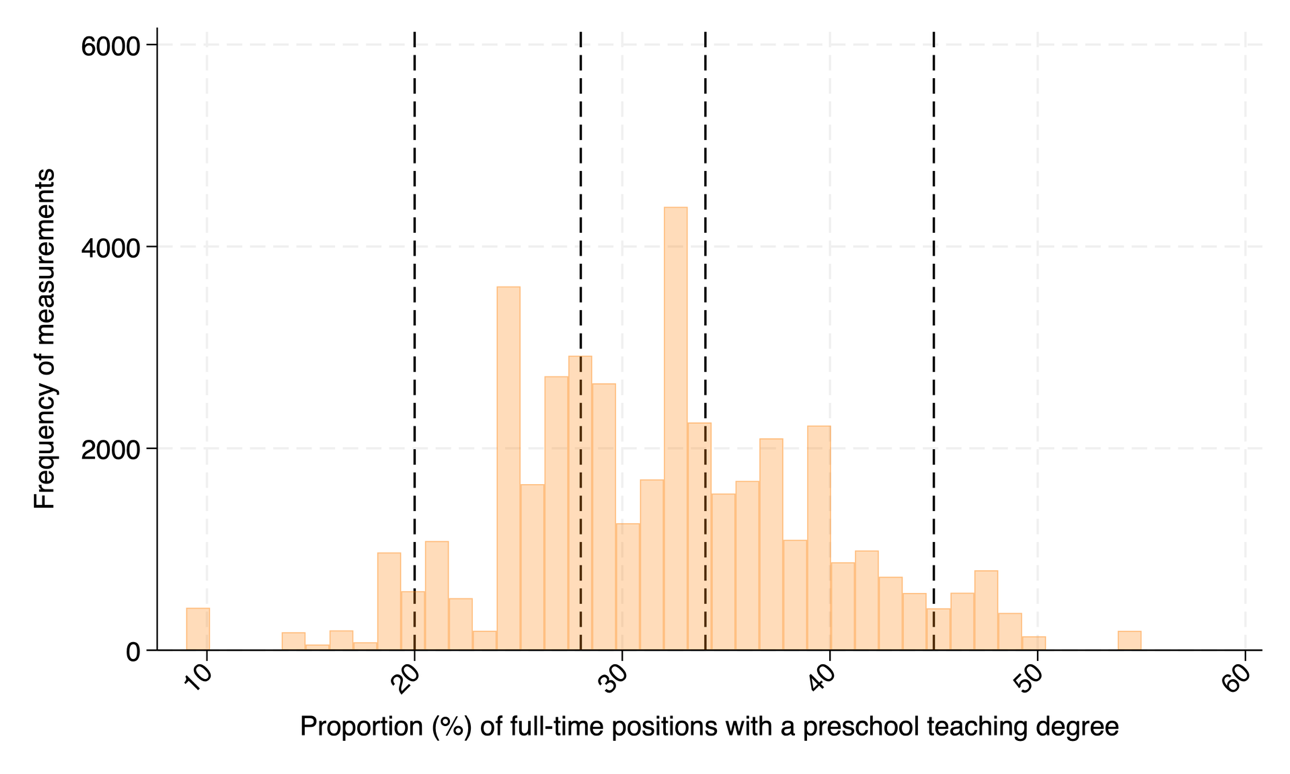
**Fig S1. A** Distribution of the proportion of full-time positions with a teaching degree using the full sample (N, measurements=43,476). Vertical lines mark the 5^th^ (20), 35^th^ (28), 65^th^ (34), 75^th^ (37), and 95^th^ (45) percentiles of the exposure.

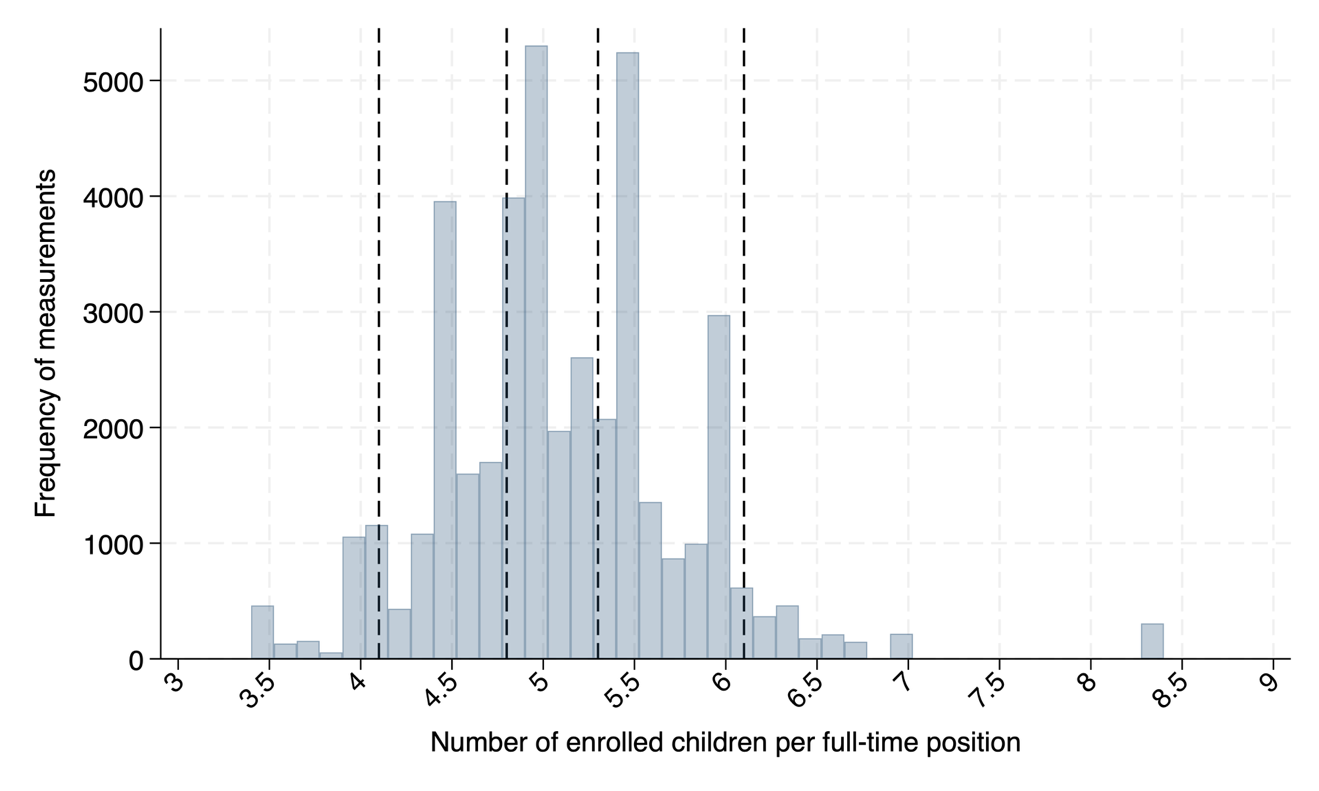

**Fig S1. B** Distribution of the number of enrolled children per full-time position (N, measurements=43,476). Vertical lines mark the 5^th^ (4.1), 35^th^ (4.8), 65^th^ (5.3), and 95^th^ (6.1) percentiles of the exposure.

| **Table S1.** Exposure values at the 25^th^, 50^th^, and 75^th^ percentiles for both exposures across all samples, along with their corresponding exposure group allocations. | | | | | | | | | |
| --- | --- | --- | --- | --- | --- | --- | --- | --- | --- |
| Teacher education* | | | | | | | | | |
| Percentile | Group allocation |  | Full sample |  | Accelerometry sample |  | SDQ sample |  | BMI sample |
| 25^th^ | Low |  | 27 |  | 27 |  | 27 |  | 27 |
| 50^th^ | Medium |  | 32 |  | 32 |  | 31 |  | 31 |
| 75^th^ | High |  | 37 |  | 37 |  | 37 |  | 37 |
| Child-teacher ratio** | | | | | | | | | |
| Percentile | Group allocation |  | Full sample |  | Accelerometry sample |  | SDQ sample |  | BMI sample |
| 25^th^ | Low |  | 4.7 |  | 4.7 |  | 4.7 |  | 4.7 |
| 50^th^ | Medium |  | 5.0 |  | 5.0 |  | 5.1 |  | 5.0 |
| 75^th^ | High |  | 5.5 |  | 5.5 |  | 5.5 |  | 5.5 |
| *Defined as the proportion (%) of full-time positions with a preschool teaching degree. **Defined as the number of enrolled children per full-time position. | | | | | | | | | |

| **Table 2.** Absolute means and relative differences (reference: 50^th^ percentile) with 95% confidence intervals derived from the restricted cubic spline models for all outcomes at the 25^th^, 50^th^, and 75^th^ percentiles of exposures. | | | | | | | | | | |
| --- | --- | --- | --- | --- | --- | --- | --- | --- | --- | --- |
| Proportion (%) of full-time positions with a preschool teaching degree | | | | |  | Number of enrolled children per full-time position | | | | |
| Reaching MVPA recommendations* | | | | | | | | | | |
| N=32,528 | | | | | | | | | | |
|  | Prevalence | 95% CI | OR | 95% CI |  |  | Prevalence | 95% CI | OR | 95% CI |
| High | 0.42 | 0.41,0.44 | 0.98 | 0.96,1.01 |  | Low | 0.43 | 0.41,0.45 | 0.99 | 0.97,1.01 |
| Medium | 0.43 | 0.42,0.45 | ref. | ref. |  | Medium | 0.43 | 0.42,0.45 | ref. | ref. |
| Low | 0.44 | 0.42,0.45 | 1.01 | 0.98,1.04 |  | High | 0.43 | 0.42,0.45 | 1.00 | 0.97,1.03 |
| Minutes MVPA* | | | | | | | | | | |
| N=32,528 | | | | | | | | | | |
|  | Minutes | 95% CI | Minutes | 95% CI |  |  | Minutes | 95% CI | Minutes | 95% CI |
| High | 60.83 | 59.62,62.04 | -0.22 | -0.84,0.40 |  | Low | 61.28 | 60.10,62.45 | -0.08 | -0.49,0.34 |
| Medium | 61.05 | 59.97,62.13 | ref. | ref. |  | Medium | 61.36 | 60.26,62.45 | ref. | ref. |
| Low | 61.01 | 59.78,62.23 | -0.04 | -0.77,0.69 |  | High | 60.91 | 59.73,62.09 | -0.45 | -1.17,0.28 |
| Minutes inactivity* | | | | | | | | | | |
| N=32,528 | | | | | | | | | | |
|  | Minutes | 95% CI | Minutes | 95% CI |  |  | Minutes | 95% CI | Minutes | 95% CI |
| High | 434.04 | 429.73,438.35 | 3.19 | 0.67,5.70 |  | Low | 431.85 | 427.68,436.03 | 0.65 | -1.09,2.38 |
| Medium | 430.85 | 427.16,434.55 | ref. | ref. |  | Medium | 431.21 | 427.44,434.98 | ref. | ref. |
| Low | 426.82 | 422.46,431.18 | -4.04 | -7.05,-1.02 |  | High | 429.43 | 425.24,433.62 | -1.78 | -4.84,1.29 |
| SDQ z-score* | | | | | | | | | | |
| N=4,385 | | | | | | | | | | |
|  | Z-score | 95% CI | Z-score | 95% CI |  |  | Z-score | 95% CI | Z-score | 95% CI |
| High | 0.01 | -0.05,0.07 | -0.02 | -0.05,0.02 |  | Low | 0.03 | -0.02,0.09 | 0.01 | -0.01,0.03 |
| Medium | 0.03 | -0.02,0.07 | ref. | ref. |  | Medium | 0.02 | -0.03,0.07 | ref. | ref. |
| Low | 0.06 | -0.00,0.11 | 0.03 | -0.01,0.07 |  | High | 0.02 | -0.03,0.08 | -0.00 | -0.04,0.04 |
| BMI z-score* | | | | | | | | | | |
| N=6,145 | | | | | | | | | | |
|  | Z-score | 95% CI | Z-score | 95% CI |  |  | Z-score | 95% CI | Z-score | 95% CI |
| High | -0.04 | -0.09,0.01 | -0.05 | -0.07,-0.02 |  | Low | -0.04 | -0.10,0.01 | -0.02 | -0.03,0.00 |
| Medium | 0.01 | -0.04,0.06 | ref. | ref. |  | Medium | -0.03 | -0.08,0.02 | ref. | ref. |
| Low | 0.06 | 0.00,0.11 | 0.05 | 0.02,0.07 |  | High | 0.03 | -0.02,0.08 | 0.06 | 0.03,0.09 |
| *Adjustments were made for child age, sex, measurement point (autumn 2020 spring 2021) and neighborhood socioeconomic index. Models including teacher education were adjusted for child–teacher ratio and vice versa. | | | | | | | | | | |

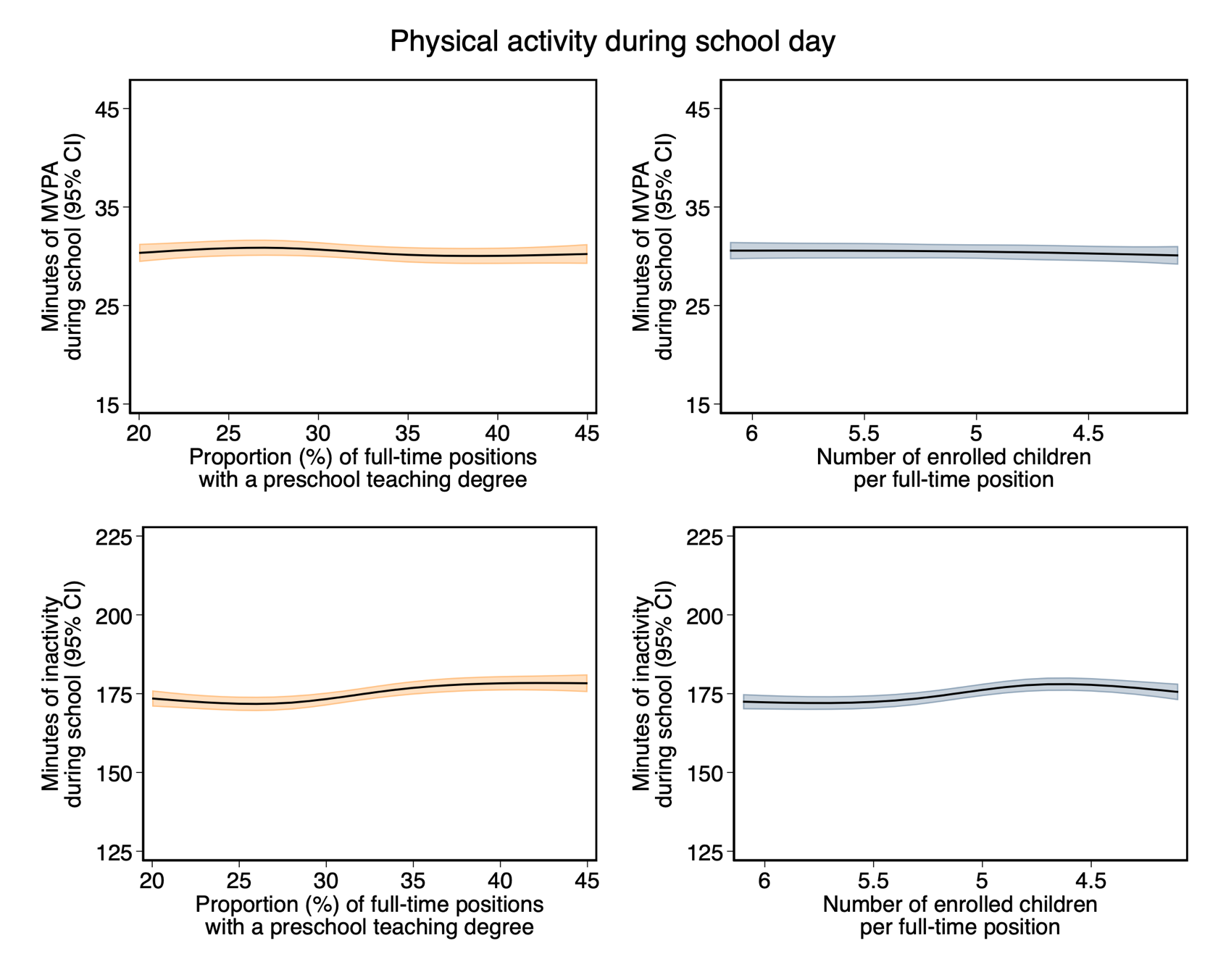

**Fig S2.** Restricted cubic spline models displaying the associations between both exposures and physical activity outcomes, using only preschool time. Knots were placed at the 5^th^, 35^th^, 65^th^ and 95^th^ percentiles, and the splines were restricted to the range between the 5^th^ and 95^th^ percentiles (Supplementary material, fig. S1 A & B). Adjustments were made for child age, sex, measurement point (autumn 2020 spring 2021) and neighborhood socioeconomic index. Models including teacher education were adjusted for child–teacher ratio and vice versa.

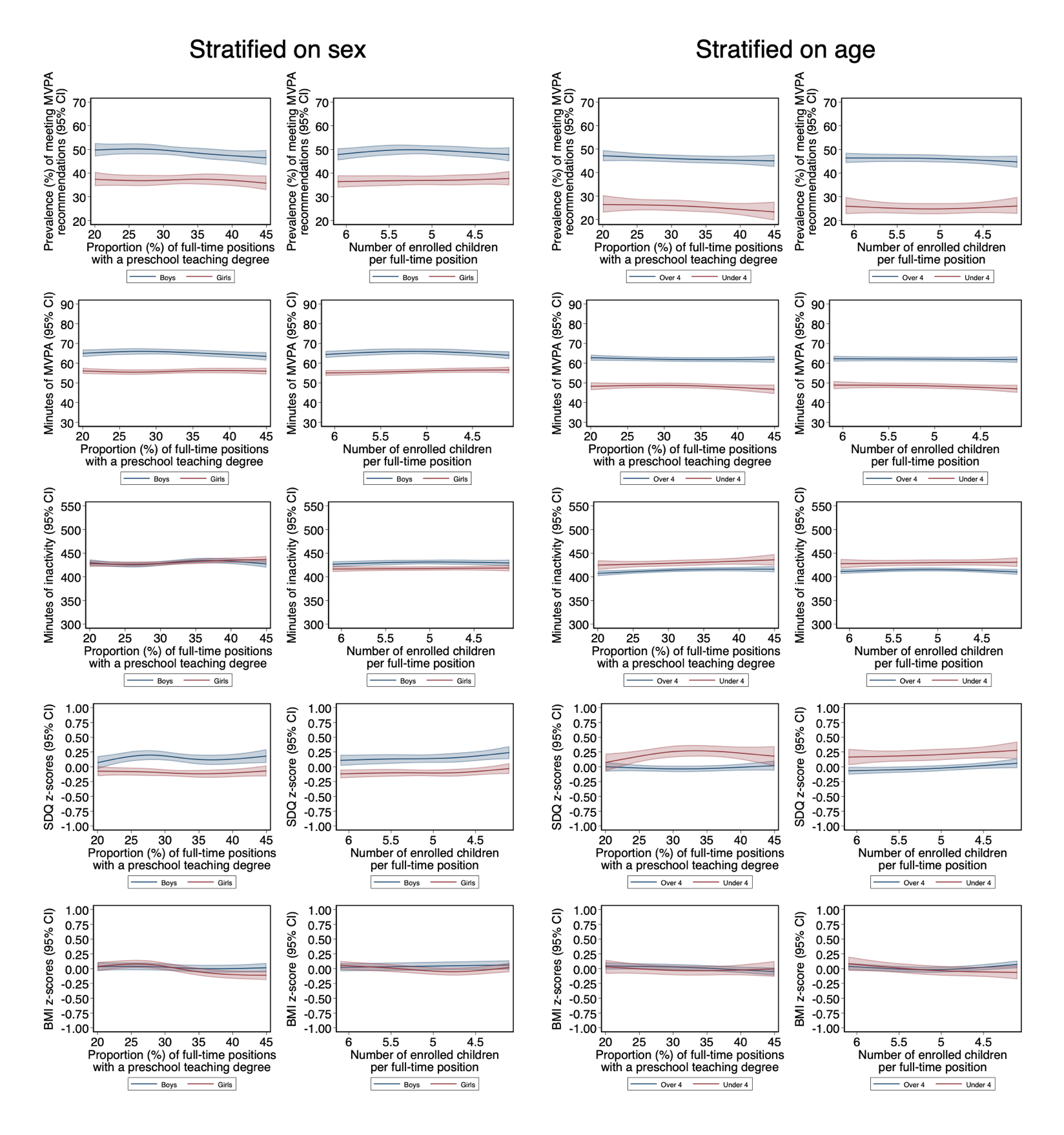

**Fig S3.** Restricted cubic spline models displaying the associations between both exposures and all outcomes. Stratifications were made by sex and age, <4 vs. ≥4 years. Knots were placed at the 5^th^, 35^th^, 65^th^ and 95^th^ percentiles, and the splines were restricted to the range between the 5^th^ and 95^th^ percentiles (Supplementary material, fig. S1 A & B). Adjustments were made for measurement point (autumn 2020 spring 2021) and neighborhood socioeconomic index. Further, the models stratified by age were adjusted for sex, whereas the models stratified by sex were adjusted for age. Models including teacher education were adjusted for child–teacher ratio and vice versa.

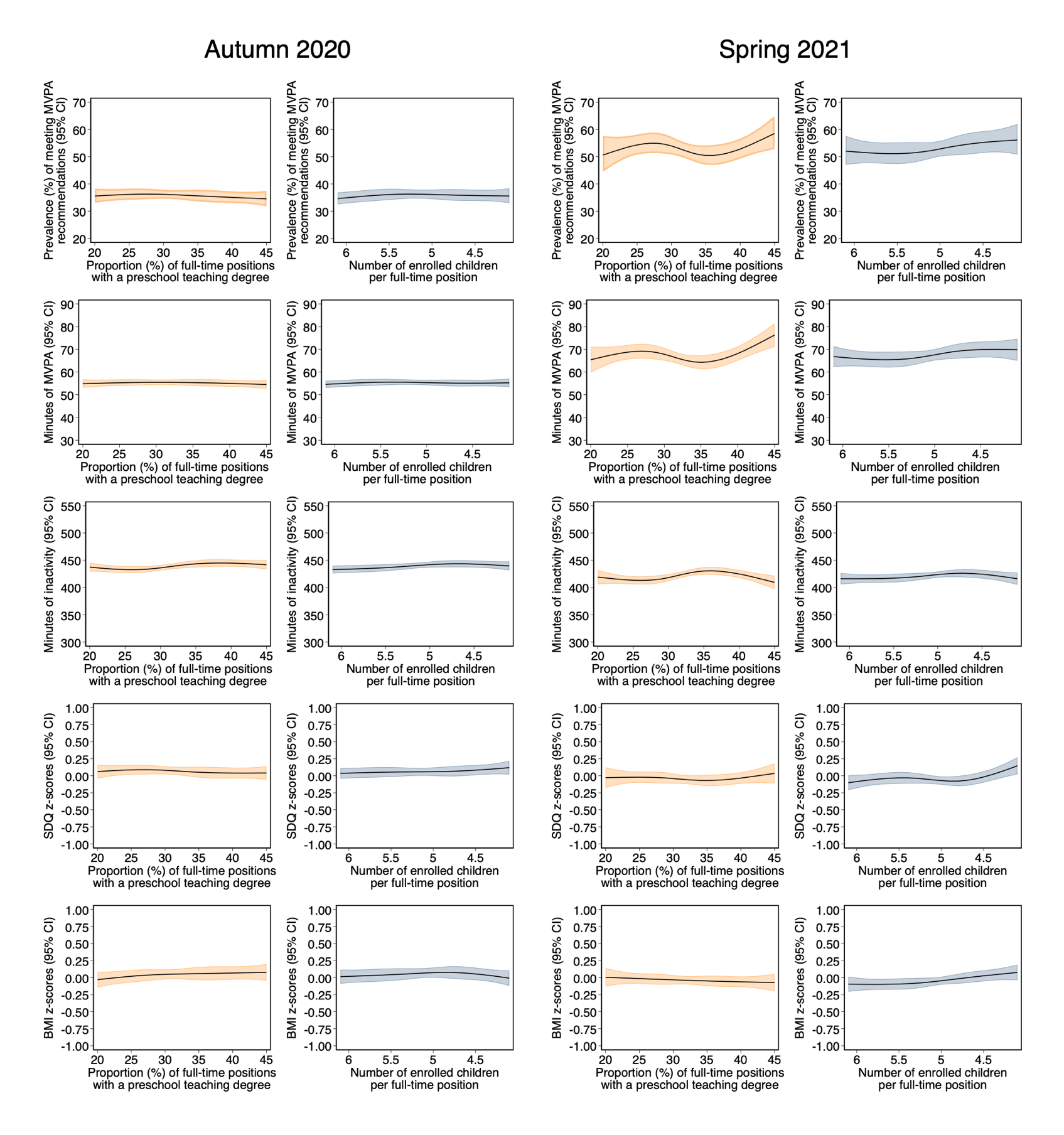

**Fig S4.** Restricted cubic spline models displaying the associations between both exposures and all outcomes, stratified on time of measurement (autumn 2020 vs. spring 2021). Knots were placed at the 5^th^, 35^th^, 65^th^ and 95^th^ percentiles, and the splines were restricted to the range between the 5^th^ and 95^th^ percentiles (Supplementary material, fig. S1 A & B). Adjustments were made for child age, sex and neighborhood socioeconomic index. Models including teacher education were adjusted for child–teacher ratio and vice versa.

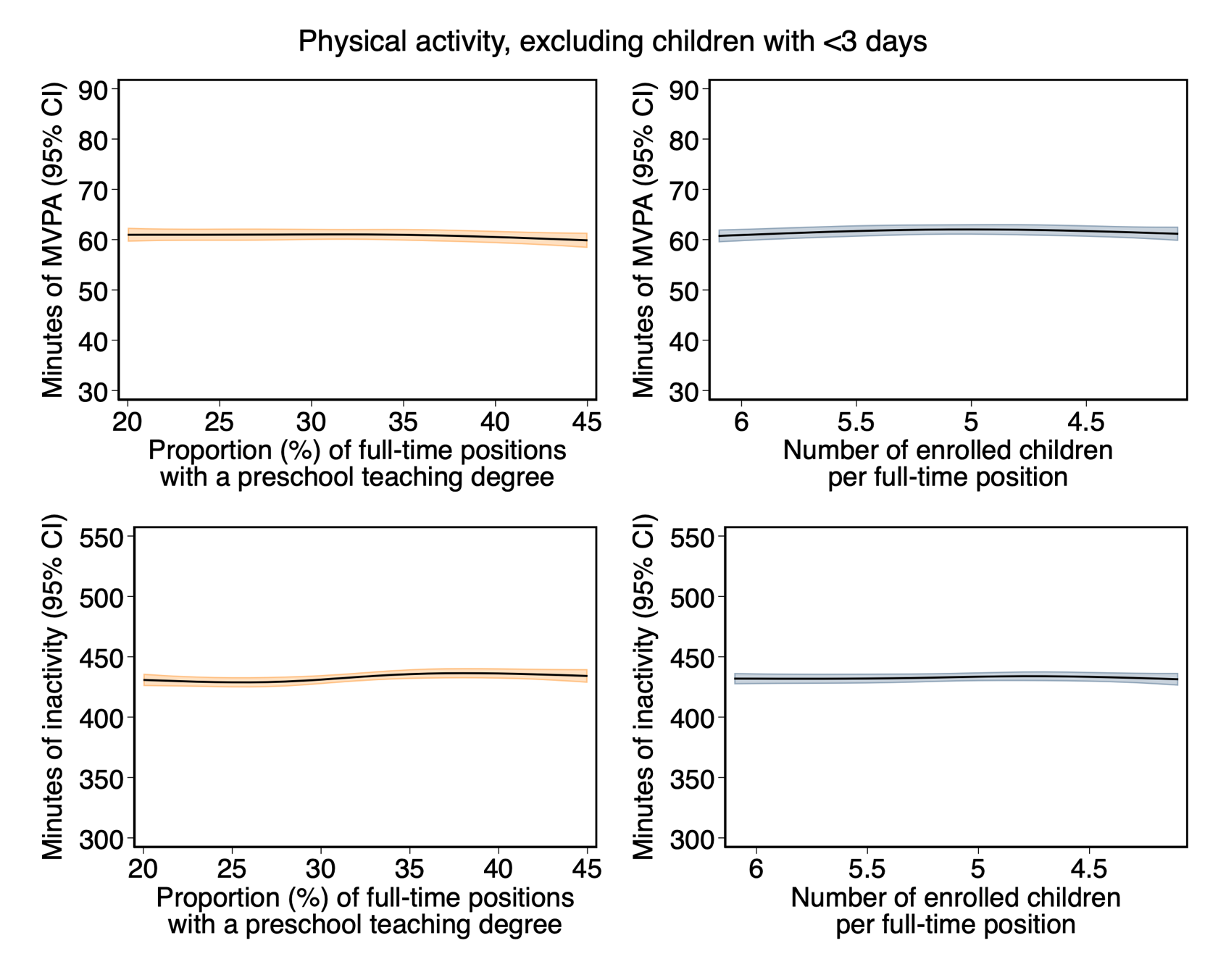

**Fig S5.** Restricted cubic spline models displaying the associations between both exposures and physical activity outcomes, including only children with 3 or more valid accelerometer days. Knots were placed at the 5^th^, 35^th^, 65^th^ and 95^th^ percentiles, and the splines were restricted to the range between the 5^th^ and 95^th^ percentiles (Supplementary material, fig. S1 A & B). Adjustments were made for child age, sex, measurement point (autumn 2020 spring 2021) and neighborhood socioeconomic index. Models including teacher education were adjusted for child–teacher ratio and vice versa.
